# Computational Biology Exposed a Common Pathogenic Mechanism in Influenza A and Guillain-Barré Syndrome

**DOI:** 10.1101/2025.08.17.25333871

**Authors:** Ye Deng, Zhongwen Tang, Qiong Chen, Xujuan Hu, Ying Jie Zhang, Meng Chen, Qiongya Wang, Fuli Ren

## Abstract

Influenza A (H1N1) is an acute respiratory infection, while Guillain-Barré syndrome (GBS) is an autoimmune peripheral neuropathy that can occur as a post-infectious complication. Clinical evidence suggests a potential link between H1N1 infection and subsequent GBS development, implying common immunopathological mechanisms. To explore this, we integrated bioinformatics and systems biology approaches to analyze transcriptome data from H1N1 and GBS patients in the GEO database, and 32 common differentially expressed genes (DEGs) were identified. Subsequent Gene Ontology (GO) and Kyoto Encyclopedia of Genes and Genomes (KEGG) pathway enrichment analyses revealed key functional associations for these DEGs. Protein-protein interaction (PPI) network analysis highlighted TLR4, TNF, and ITGAM as central hub genes, uncovering potential shared molecular pathways between H1N1 and GBS. Furthermore, analysis of hub gene interactions with microRNAs (miRNAs), transcription factors (TFs), and related diseases facilitated the prediction of potential therapeutic drugs. Molecular docking simulations were performed to validate predicted drug interactions with the hub gene products. Collectively, these findings delineate shared molecular mechanisms and provide insights for targeted therapy in patients developing GBS post-H1N1 infection.

## Introduction

Influenza A is an acute respiratory infection caused by influenza A viruses, and its global epidemic infects millions of people each year, with significant morbidity and mortality.[1, 2] H1N1, the most prevalent subtype of the influenza A virus, has been demonstrated to induce the release of cytokines and trigger an inflammatory response through the activation of both innate and adaptive immune responses in the host, resulting in respiratory symptoms and systemic complications[3]. Guillain-Barré syndrome (GBS) is a post-infectious autoimmune peripheral neuropathy characterised by acute flaccid paralysis. This condition is associated with molecular mimicry or a cross-immune response, which causes the immune system to attack the myelin sheath or axons of peripheral nerves[4]. A substantial body of research in the field of epidemiology has demonstrated that between 20 and 30 percent of patients diagnosed with Guillain-Barré syndrome (GBS) have experienced a secondary onset of the condition following a viral or bacterial infection. Among the various causative agents identified, influenza virus has been identified as a significant trigger for this secondary manifestation. For instance, during the 2009 influenza A H1N1 pandemic, the incidence of Guillain-Barré syndrome (GBS) increased significantly, further suggesting a potential association between the two in immunopathological mechanisms[5].

Whilst clinical observations support a temporal association between influenza and GBS, the molecular mechanism between the two remains unclear. A paucity of research has hitherto been conducted on the common pathogenic network across diseases, with the majority of studies focusing on the immune or genetic characteristics of a single disease. For instance, research has been conducted on the activation of interferon signalling pathways in influenza A or the role of anti-ganglioside antibodies in GBS. Furthermore, extant treatment strategies principally target immune abnormalities in GBS, but prophylactic or targeted interventions for post-influenza A GBS remain limited[6]. In recent years, bioinformatics and systems biology approaches have demonstrated unique advantages in the identification of molecular interaction networks in complex diseases. These approaches include the identification of core regulatory modules through the integration of multi-omics data and the utilisation of machine learning models to predict potential therapeutic targets. However, the application of such methods in the crossover study of H1N1 and GBS has not been fully explored.

The objective of this study was to investigate the co-pathogenesis of influenza A H1N1 virus infection and GBS, and to explore potential therapeutic agents. Firstly, the GEO database transcriptome datasets of H1N1 and GBS were analysed to identify the DEGs of the two diseases, respectively, and then the common DEGs were obtained by comparison. It is vital to note that, we did not use these common differentially expressed genes (DEGs) for gene ontology (GO) functional annotation and Kyoto Encyclopedia of Genes and Genomes (KEGG) pathway enrichment analysis, as the platforms of the two datasets used were different and there was a gap in data volume. In this study, GO and KEGG of the two datasets were analyzed separately. Then take the intersection according to the degree of importance. Subsequently, the PPI network was constructed, hub genes were screened out by topological analysis, and the key genes with the highest connectivity were selected for further study. In order to verify the clinical value of this key gene, the regulatory mechanism of common DEGs in H1N1 infection was revealed through transcription factor-target gene interaction network analysis. Finally, the database analysis yielded the drugs associated with the hub genes, and then used the obtained drug molecules to perform simulated docking experiments with the target protein molecules of H1N1 and GBS. The experimental workflow, from data collection to analysis, is depicted in Fig 1

**Fig 1.**
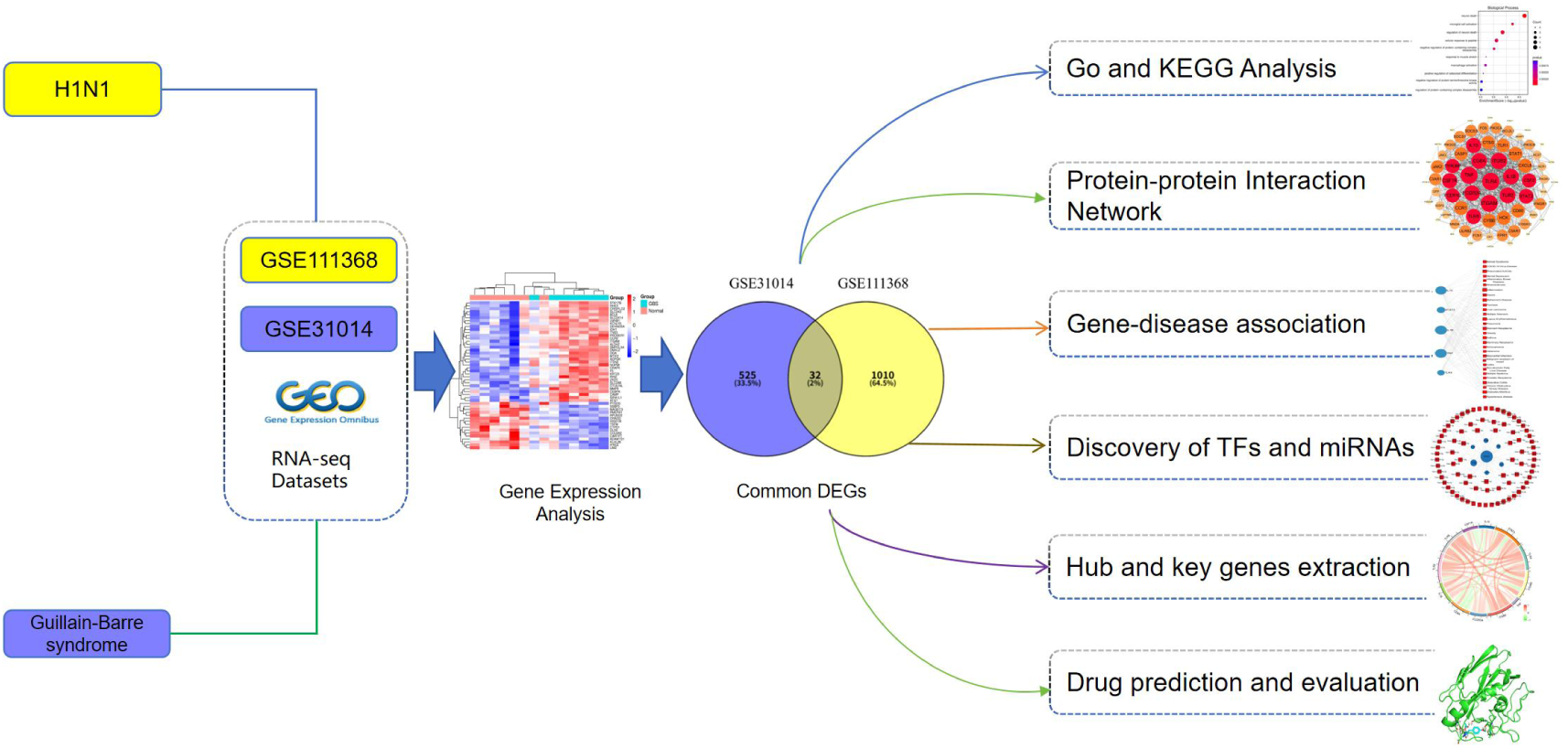
Schematic diagram of the research process in this paper.

## Materials and Methods

The GEO database (https://www.ncbi.nlm.nih.gov/geo/) was accessed, and the GSE111368 dataset was selected, which was generated on the GPL10558 microarray platform (Illumina HumanHT-12 V4.0)[7]. The dataset under consideration contains 109 samples infected with influenza A (H1N1) and 130 samples from healthy subjects. The sequencing platform utilised the Illumina Human HT-12 V4.0 expression microarray chip. The GSE31014 dataset was generated using the GPL96 platform (Affymetrix Human Genome U133A Array [HG-U133A]), comprising seven GBS patient samples and seven healthy control samples [8–9]. The identification of disease-associated DEGs was achieved through a comparative analysis of the GSE111368 (H1N1 infection) and GSE31014 (GBS) mRNA datasets. Utilising the limma R package in conjunction with empirical Bayes moderation, we implemented linear models to extract differentially expressed genes (DEGs) for each disease. The selection of statistically significant genes was undertaken utilising dual thresholds. Benjamini-Hochberg adjusted P < 0.05 was employed for false discovery rate control and absolute log2 fold-change (log2FC) ≥ 1. Finally, shared DEGs between the two datasets were identified via the VENN v2.1 online tool (https://bioinfogp.cnb.csic.es/tools/venny/) [10–11].

### Functional enrichment analysis of common DEGs

Functional enrichment analysis is a bioinformatics approach that identifies statistically overrepresented biological annotations with the aim of elucidating the potential mechanisms of DEGs. The method under scrutiny has been demonstrated to detect significant enrichment of GO terms, KEGG pathways and disease-associated signatures. This capability enables the delineation of the functional contributions of specific gene collections to particular pathophysiological processes. In this study, we conducted go and pathways analyses on H1N1 infection and GBS respectively. Among the documents obtained from the go analyses and pathways analyses of both, the top ten significant common functions and pathways were selected based on the minimum adjusted P-value as the ranking criterion to explore the common molecular mechanisms of these two diseases.

Specifically, we used GO analysis to functionally classify common DEGs, including biological processes (BP), molecular function (MF), and cellular components (CC), to clarify their roles in key pathological processes such as immune regulation and neuroinflammation. At the same time, based on KEGG pathway enrichment analysis, the signaling pathways involved in these genes (such as virus-host interaction, autoimmune response, etc.) were further analyzed, so as to further discover the potential association mechanism between H1N1 infection and the pathogenesis of GBS[11–13].

### Protein-protein interaction network analysis

Proteins execute their mechanistic roles within cellular systems through interactions in protein-protein interaction (PPI) networks. To elucidate disease-related protein functions, we constructed PPI networks using common DEGs for subsequent interaction analysis[14]. The analysis was performed using STRING (v12.0; https://www.string-db.org/), a comprehensive database of known and predicted protein associations, including physical interactions and functional linkages. Common DEGs were entered into STRING and a confidence threshold (composite score > 0.4) was applied to generate PPI networks[15, 16].

The PPI networks were imported into Cytoscape (v3.10.3) for visualization and downstream analysis. Hub genes were identified using CytoHubba, a Cytoscape plugin that calculates 12 network centrality algorithms to rank node importance[14,17]. Based on validation studies demonstrating its superior discriminative power, the Maximum Clique Centrality (MCC) method was selected to extract the twelve most significant hub genes..

### Transcriptional Regulatory Network Mapping via TF and miRNA Interactions

To decode the transcriptional regulatory architecture underlying shared DEGs in H1N1 influenza and GBS, we leveraged the NetworkAnalyst platform (https://www.networkanalyst.ca/) for multi-omics integration[18]. Firstly, we utilized the JASPAR database to identify reliable transcription factor binding sites, and at the same time combined the experimentally validated miRNA-target gene interactions data in miRTarBase and TarBase[19–21]. We constructed a comprehensive DEG-TF-miRNA regulatory network; subsequently, we used Cytoscape (v3.10.3) to perform network visualization and analysis, focusing on the screening of core transcription factors and key miRNAs that regulate multiple common DEGs, and these regulatory nodes may be involved in the common pathogenesis of the two diseases by influencing processes such as virus-host interactions and neuroimmune regulation.

### Gene-disease association analysis networks

The DisGeNET database (https://www.disgenet.org/) was utilised to construct gene-disease association networks, with molecular relationships being systematically aggregated from experimental data, GWAS repositories, and curated literature evidence [22]. The NetworkAnalyst platform was utilised to integrate these associations with multi-omics networks. This facilitated the delineation of disease mechanisms and enabled the construction of comorbidity networks between H1N1 and GBS.

### Therapeutic Compound Screening via Gene Signature Drug Repositioning

We predicted protein-drug interactions (PDIs) and identified repurposable therapeutic agents by targeting common DEGs shared between H1N1 influenza and GBS. Using the Drug Signature Database (DSigDB) embedded within the Enrichr platform, we performed gene set enrichment analysis to discover compounds significantly associated with these DEG signatures (FDR<0.05)[23]. The Enrichr repository (http://amp.pharm.mssm.edu/Enrichr) provides comprehensive, genome-scale gene set libraries for enrichment analysis, with DSigDB specifically curating drug/compound-target gene relationships accessible through its Disease/Drug Functions module[24].

### Structure-Based Virtual Screening via Molecular Docking

Molecular docking, a computational drug discovery approach predicting ligand-target binding modes, was employed to identify therapeutic compounds and establish structure-activity relationships without prior knowledge of target modulators [25]. Specifically for this study, we selected H1N1 viral targets — including receptor-binding proteins (hemagglutinin/HA, neuraminidase/NA) and functional non-receptor proteins (M2 ion channel regulating endosomal pH, nucleoprotein/NP for RNA packaging)— alongside GBS-associated targets (gangliosides, myelin proteins) through systematic literature analysis. High-resolution crystal structures were retrieved from the PDB database (via NCBI) and compound libraries from PubChem. Using the CB-Dock2 platform implementing cavity-detection guided blind docking (https://cadd.labshare.cn/cb-dock2), we performed energy minimization of protein structures with the AMBER force field prior to docking simulations. Binding affinity was quantitatively evaluated by weighted-average Binding Energy (BE) scoring, where BE ≤ -8.0 kcal/mol indicated high-affinity interactions. Finally, binding poses were analyzed in PyMOL v2.5.5 with critical intermolecular distances (H-bond/ π - π stacking < 3.5Å) measured, though molecular dynamics simulations were not conducted for conformational validation.

## Result

### Identification of common gene expression signatures between H1N1 and GBS

Severe H1N1 influenza infection has been observed to trigger GBS in susceptible individuals during the post-infection immune dysregulation phase (5 days to 3 weeks) through molecular mimicry mechanisms analogous to Campylobacter jejuni pathogenesis. However, further characterisation is required to determine the specific influenza-specific antigenic targets involved in this process. It is noteworthy that patients who developed GBS following H1N1 infection exhibited doubled mortality rates compared to classical GBS cases. This phenomenon can be attributed to the exacerbation of respiratory compromise and autonomic dysfunction.

Transcriptomic profiling of peripheral blood samples (GSE111368, H1N1; GSE31014, GBS) identified 1,042 differentially expressed genes (DEGs), with 141 being found to be upregulated and 175 downregulated in H1N1, and 40 being found to be upregulated and 517 downregulated in GBS. Volcano plots illustrated prominent expression shifts in both conditions (Fig. 2A-B), while hierarchical clustering heatmaps revealed distinct disease-specific signatures (Fig. 2C-D). Of particular significance was the observation that the 12 most significantly dysregulated H1N1-associated DEGs converged on neuroinflammatory pathways and immune response regulation, suggesting shared mechanistic underpinnings. Venn analysis (Fig. 2E) delineated 32 conserved DEGs between the two conditions. A comprehensive investigation into the functional annotation of these overlapping signatures has revealed that there are some commonalities in the functional mechanisms and interactions between H1N1 and GBS.

**Fig 2.**
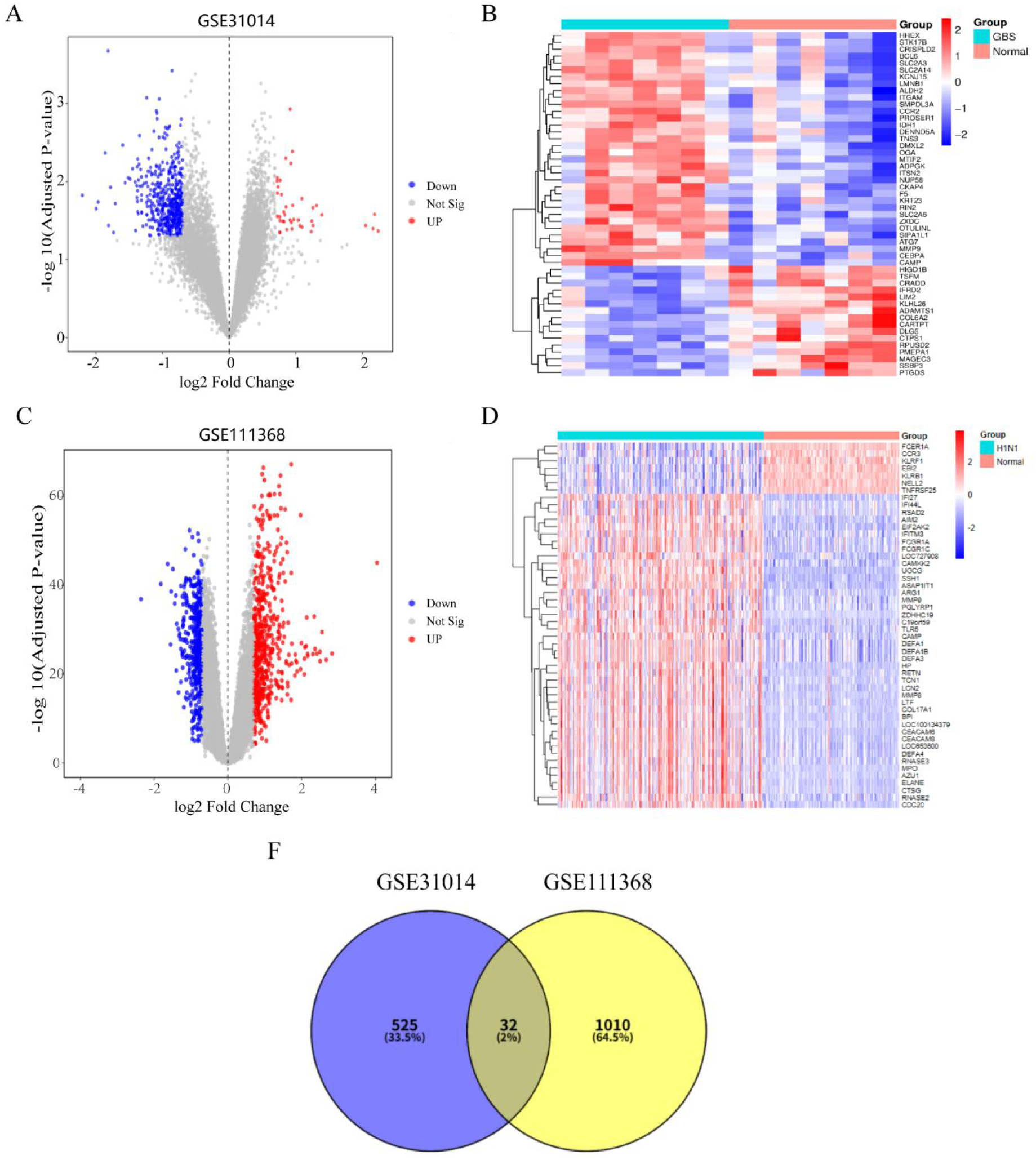
Differential gene expression profiling in Guillain-Barré syndrome (GBS) and H1N1 influenza datasets. (A) Volcano plot of DEGs in GBS dataset (red: up-regulated; blue: down-regulated; gray: non-significant genes; P<0.05). (B) Heatmap clustering analysis of top 50 DEGs in GBS. (C) Volcano plot of DEGs in H1N1 dataset. (D) Heatmap clustering analysis of top 50 DEGs in H1N1. (F) Venn diagram of common DEGs between GSE31014 and GSE111368 datasets.

### Functional enrichment analysis of differentially expressed genes

As illustrated in Fig 3A-B, bubble plots are employed to present the results of gene ontology (GO) enrichment analysis, which is categorised into three distinct groups. The following three categories are to be considered: biological process (BP), cellular component (CC), and molecular function (MF). In these visualisations, colour depth corresponds to statistical significance (-log10(p-value)), while the size of each bubble reflects the number of genes in each term. Specifically, the results of the BP in Fig 3A analysis revealed highly enriched immune responses: Items such as Gram-positive bacterial defense response, antibacterial humoral response, and viral defense response are red in color and have a large number of genes, indicating a significant enrichment of immune defense-related genes. The research subjects may be in a state of pathogen stimulation and immune activation. Fig 3B presents a multi-dimensional functional analysis of the genes based on GO enrichment. In the biological process (BP) category, the enrichment of immune defense response pathways confirms their central role in the immune response, indicating that the research subjects may be under pathogenic stimulation or in a state of immune activation. Both graphs point to "significant activation of the immune defense response" and the two figures from different visualization forms jointly verify that the immune defense response is the core functional enrichment direction of the research object.

**Fig 3.**
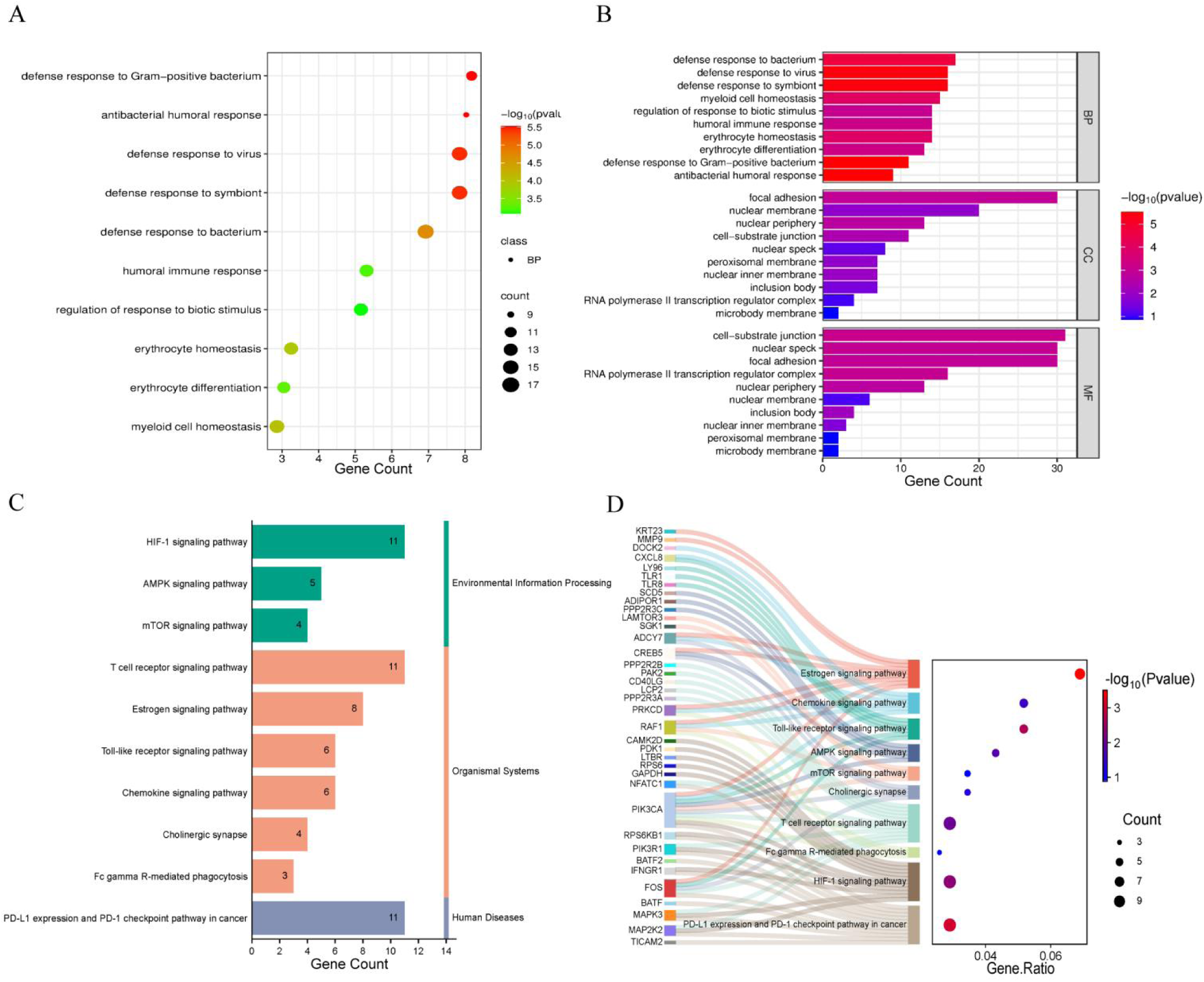
The significance analysis results obtained by analyzing the gene Ontology (GO) and KEGG evaluations of the two datasets. (A) Scatter plot visualization of BP terms significantly enriched by GO analysis. (B) The bar chart of Gene Ontology (GO) enrichment analysis divided into BP (Biological Process), CC (Cellular Component), MF (Molecular Function). (C) Bar chart depicting the results of KEGG pathway enrichment analysis for the overlapping pathways identified in the two datasets.. (D) Sankey diagram visualizing the enrichment relationships of shared KEGG pathways between the two datasets.

The Fig 3C-D presents a visual synthesis of KEGG signaling pathway enrichment analysis results and gene-pathway association profiles. In Fig 3C, results demonstrate significant activation of immune response, hypoxic adaptation, and tumor immunity-associated pathways (FDR < 0.05), suggesting that immune activation, a hypoxic microenvironment, or a tumor-associated state aligns with the clinical symptoms of influenza and GBS patients. In Fig 3D, 37 genes, including KRT23, MMP9, CXCL8, TLR8, PIK3CA, MAPK3, etc., were mapped to 10 core signaling pathways through Sankey junction lines. Among them, Toll-like receptor signaling pathways involving LY96, TLR8, TICAM2, chemokine signaling pathways enriched CXCL8 and DOCK2, and T cell receptor signaling pathways CD40LG, LCP2, and NFATC1 showed high correlations. In particular, the simultaneous enrichment of PD-L1/PD-1 cancer immune checkpoint pathways such as PIK3CA and PIK3R1 and AMPK/mTOR metabolic pathways such as PDK1 and RPS6KB1 suggests a possible synergistic effect between metabolic reprogramming and immune escape mechanisms. Collectively, these findings integrate across Fig 3A-D to identify functional convergence in neurodegenerative processes and anti-viral immune responses through coordinated dysregulation of shared gene networks.

### Protein Interaction Network Analysis Reveals Core Molecular Drivers

A protein-protein interaction (PPI) network was constructed using the 32 common differentially expressed genes (DEGs) that were identified as being differentially expressed between H1N1 influenza and Guillain-Barré syndrome (GBS). The network under consideration (Fig. 4A) was constructed utilising the STRING database and subsequently visualised in Cytoscape v3.10.3. The network comprises 73 nodes interconnected by 770 edges, with the size and colour intensity of the nodes correlating with their interaction centrality. It can be observed that larger, darker nodes indicate greater functional significance. A comprehensive topological analysis was conducted using five distinct algorithms (MCC, Degree, MCN, Closeness, DMNC) with CytoHubba, which resulted in the identification of pivotal regulatory genes. The Maximal Clique Centrality (MCC) method was employed to identify 32 genes of particular significance in the context of the study. Convergence across all algorithms led to the identification of 10 core overlapping regulators: TLR4, ITGAM, TNF, FCGR3A, CD8A, IL1B, TLR2, TLR8, CSF1R, IL10, and STAT3 (see Table 1 for complete rankings). Submodule analysis in Fig. 4B demonstrates tight functional clustering among these hub genes, with TLR4—a pathogen recognition receptor critical for innate immune activation (NCBI Gene:7099)—emerging as the highest-connectivity node.

**Fig 4.**
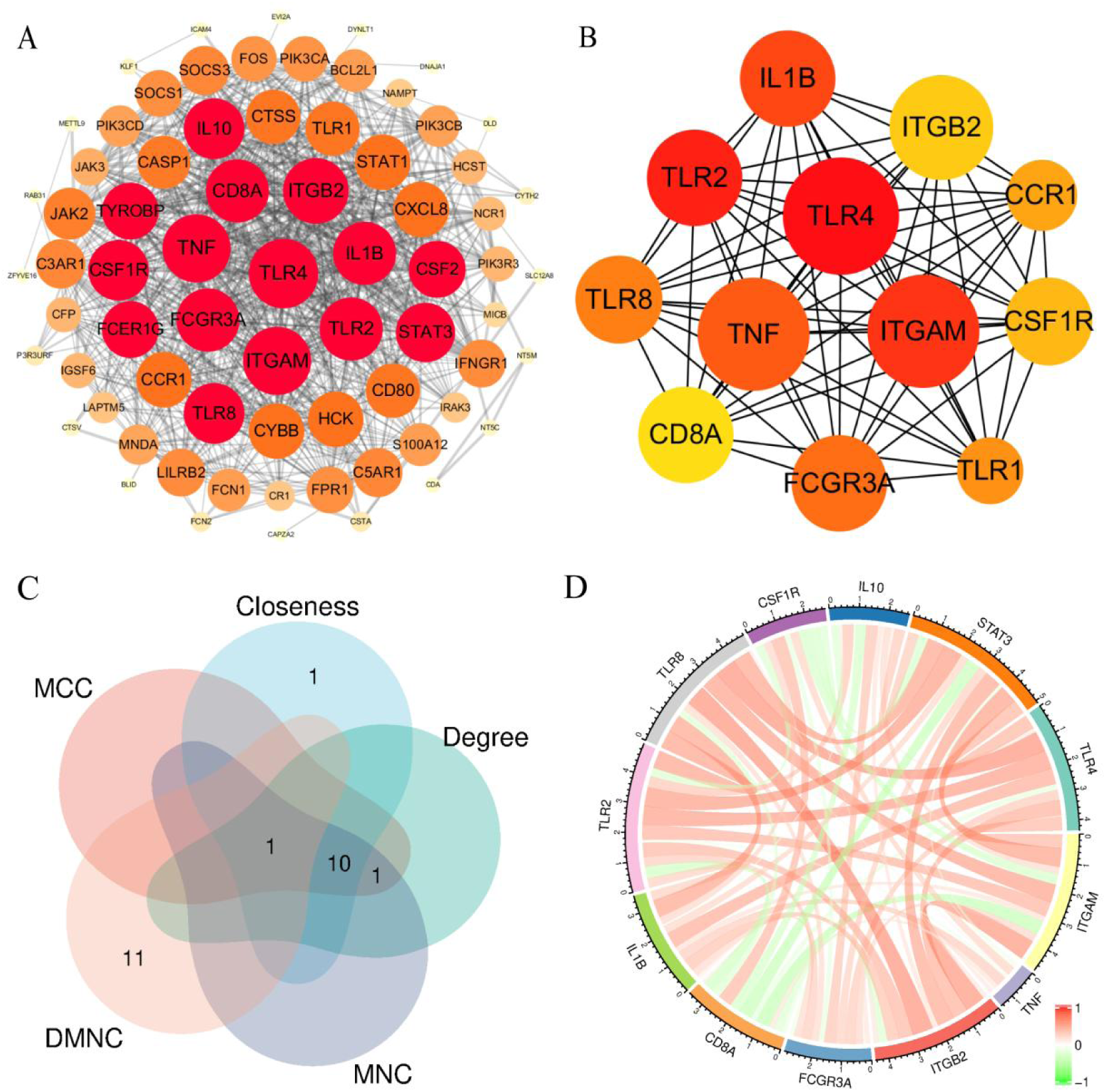
Hub gene identification and interaction analysis. (A) Protein-protein interaction network of 32 shared differentially expressed genes (DEGs) visualized in Cytoscape. (B) Key functional clusters identified through MCC algorithm screening. (C) Intersection of hub genes derived from five distinct computational methods (Venn diagram). (D) Co-expression network of the top 12 hub genes ranked by MCC algorithm scores.

**Table 1.**
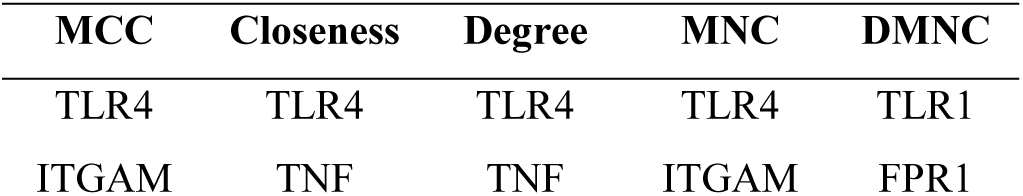
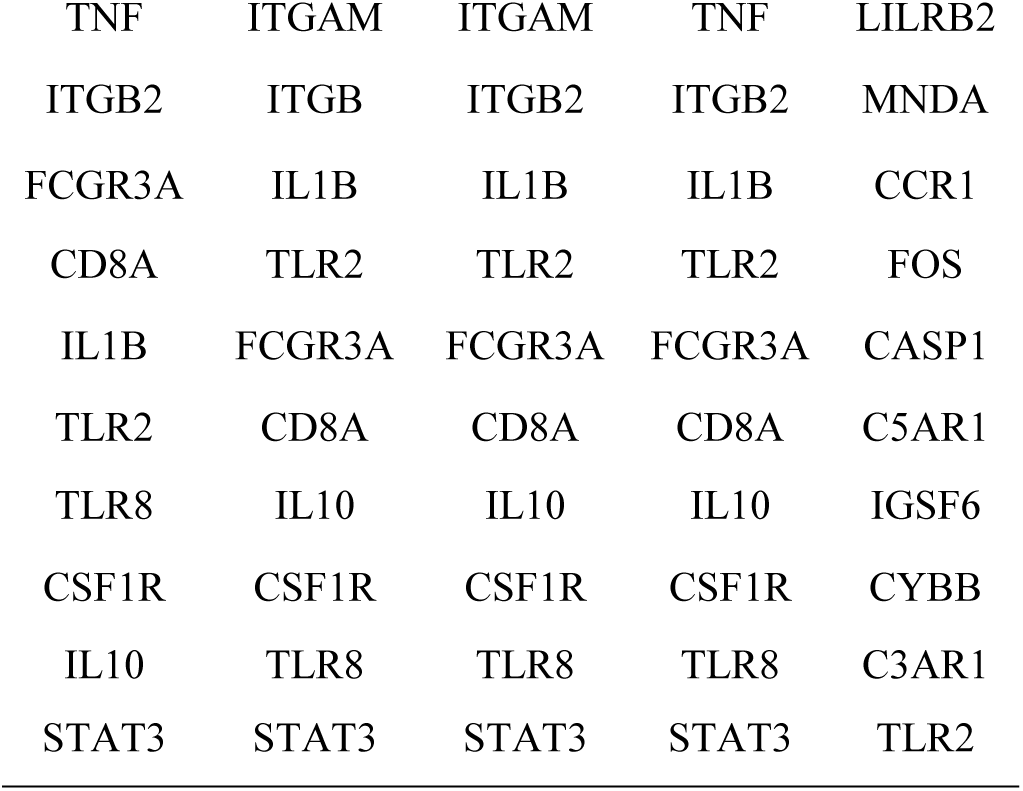
CytoHubba’s AlphaMc using five algorithms (MCC, Closeness, Degree, MNC, and DMNC)

Further characterisation included the creation of a Venn diagram (Fig. 4C), which confirmed a high level of concordance between the algorithms in terms of Degree centrality values. Additionally, gene co-expression networks were constructed via gene co-expression network analysis (GCNA) for the top 12 ranked genes (Fig. 4D). The integrated results of this study establish that TLR-mediated immune activation and cytokine signalling pathways are central mechanisms that link viral infection and the pathogenesis of autoimmune neuropathy.

### Differential expression analysis of GBS-H1N1 common DEGs

The expression profiling of key genes across H1N1 influenza and GBS datasets revealed significant upregulation of TLR4 in GBS samples relative to controls (Figs. 5A-B). This finding is consistent with the established role of TLR4 in pathogen recognition via LPS sensing and innate immune activation. A correlation analysis of shared differentially expressed genes was conducted, which identified TLR8 and ITGB2 as exhibiting the strongest positive relationship (r=0.78, Fig. 5C). This finding suggests that TLR8-mediated viral RNA recognition may upregulate ITGB2, thereby enhancing immune cell adhesion and migration during the process of pathogen clearance. The finding is mechanistically reinforced by the co-regulation of TLR4 and TLR8 (r=0.76), indicating synergistic activation during co-infections that amplifies production of pro-inflammatory cytokines, including TNF-α and IL-6. As illustrated in Figure 5D, ITGB2 demonstrates predominant expression in both disease contexts. In the context of H1N1 infection, this phenomenon may be indicative of viral activation of TLR3/7/8 and RIG-I pathways in myeloid cells, thereby promoting the formation of the ITGB2-integrin complex (CD11b/CD11a), which in turn facilitates the trafficking of immune cells. In contrast, elevated ITGB2 levels in GBS are associated with anti-ganglioside antibody-mediated complement activation. This process involves the recruitment of ITGB2-expressing macrophages and T-cells to peripheral nerves, where they induce demyelination and axonal damage. This correlation has been further substantiated in experimental autoimmune neuritis models, which demonstrate that ITGB2 expression levels are directly proportional to the severity of neuroinflammation. The integrated findings establish a continuum wherein viral-triggered immune mechanisms converge with autoimmune processes through dysregulation of shared molecular pathways involving TLR signalling and integrin-mediated inflammation.

**Fig 5.**
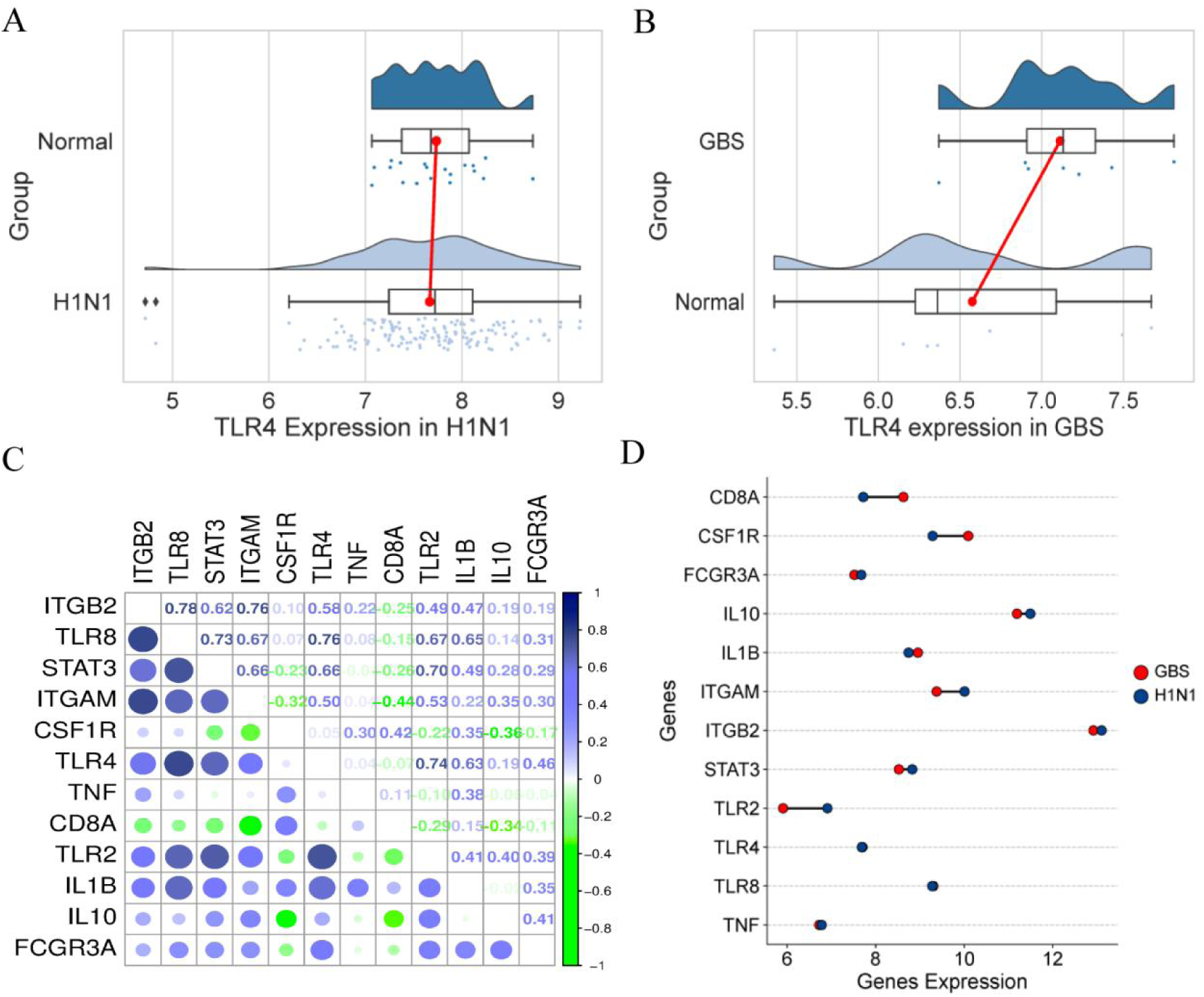
Expression patterns and correlations of key genes in H1N1 and Guillain-Barré syndrome (GBS) datasets. (A-B) Differential expression of TLR4 in H1N1 (A) and GBS (B) datasets. (C) Correlation matrix of significantly DEGs. (D) Comparative expression profiling of key DEGs across H1N1 and GBS datasets.

### Transcriptional and Post-transcriptional Regulatory Networks of Core H1N1-GBS Genes

In order to elucidate the transcriptional-level regulation of common differentially expressed genes (DEGs), integrated regulatory networks were constructed, mapping transcription factors (TFs) and microRNAs (miRNAs) through TarBase and miRTarBase databases. As illustrated in Figures 6A-B, the DEG-TF interaction network is visualised, whereby circular nodes represent DEGs and square nodes denote TFs. The size of each node is scaled by its connectivity degree, with larger nodes indicating critical network hubs. Key DEGs with high connectivity (TLR4, STAT3, TNF, ITGAM, IL-10) demonstrate multifaceted roles; a. TLR4 (a key innate immune receptor) detects pathogens and modulates inflammatory responses during H1N1 infection, while its elevated expression in GBS patients correlates with disease severity; b. STAT3 compensated for H1N1 NS1-mediated STAT1 inhibition, thereby maintaining antiviral defences. Furthermore, the research revealed that STAT3 inhibitors attenuated GBS-like neuroinflammation in animal models [26]. Among the transcription factors (TFs) analysed, NF-κB, JUN, SP1, GATA3, and HSF1 exhibited the highest regulatory significance..

**Fig 6.**
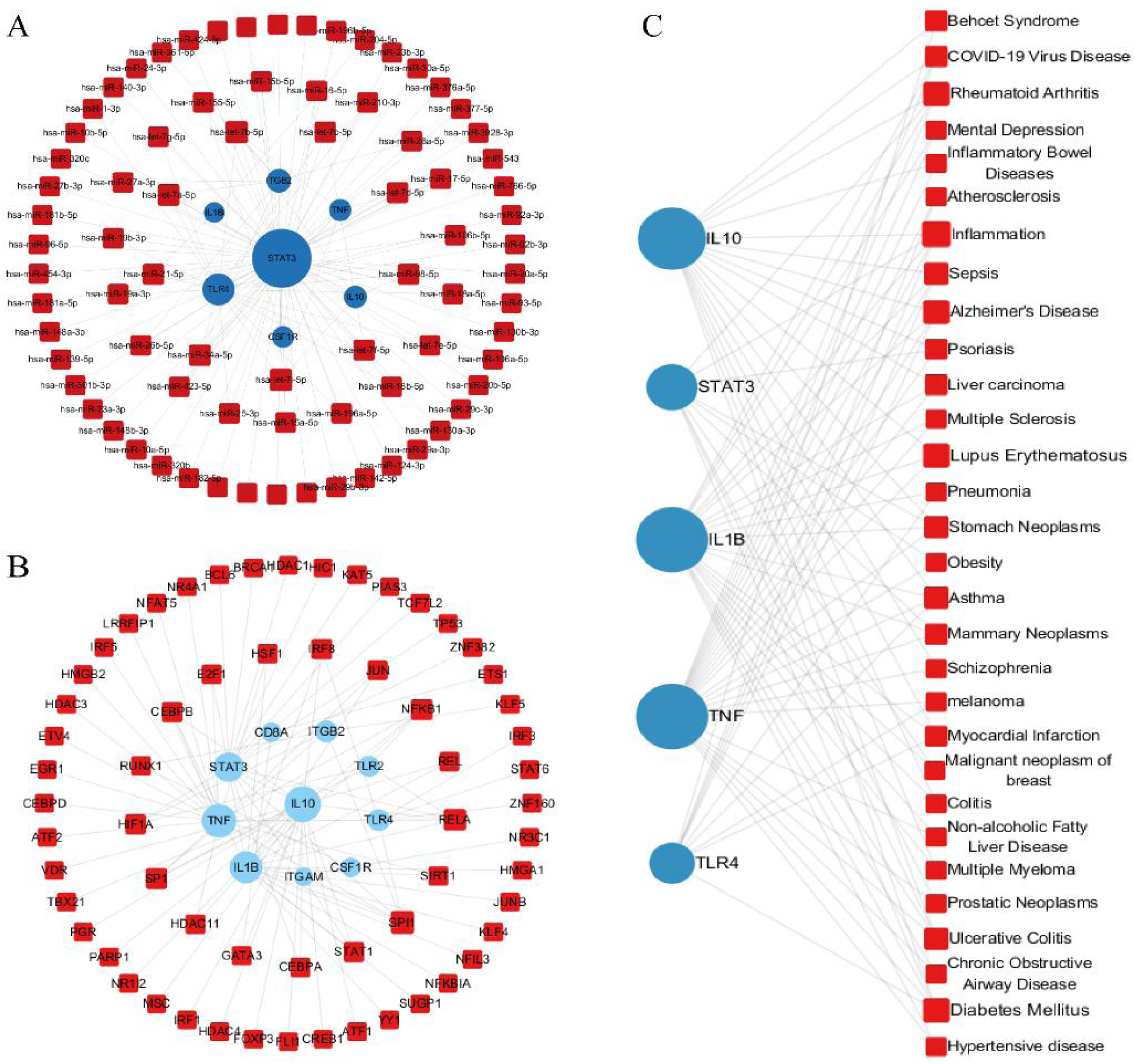
Regulatory networks illustrating interactions among DEGs, miRNAs, transcription factors (TFs), and diseases. (A) DEGs-miRNA regulatory network: Blue circular nodes represent differentially expressed genes (DEGs); red square nodes represent miRNAs. (B) DEGs-TF interaction network: Blue circular nodes indicate DEGs; red square nodes denote transcription factors. (C) Gene-disease association network: Blue circular nodes show DEGs; red square nodes represent disease entities.

Complementary networks of differentially expressed genes (DEGs) and microRNA (miRNA) (Fig. 6C) identified seven genes (STAT3, IL-1 β, IL-10, TLR4, TNF, ITGB2, CSF1R) with the strongest interactions between DEGs and miRNAs. The hub miRNAs included hsa-miR-34a-5p, hsa-miR-98-5p, hsa-miR-19a-3p, hsa-miR-27a-3p, and hsa-miR-25a-3p. It is of crucial importance to note that hsa-miR-34a-5p has been observed to exhibit dual pathological roles. Firstly, in cases of H1N1, it has been demonstrated to attenuate cytokine storms by inhibiting NF-κ B. In Guillain-Barré syndrome (GBS), it has been demonstrated that this exacerbates autoimmunity by promoting Th17/M1 polarization, which has been shown to positively correlate with disease severity [27]. This suggests that its inhibition could promote neural repair.

### Therapeutic Discovery Validated by Multi-Target Molecular Docking

Our molecular docking strategy targeted pathophysiologically critical proteins: H1N1 surface glycoproteins hemagglutinin (HA) and neuraminidase (NA) mediating viral-host adhesion [38], alongside GBS autoantigen myelin protein zero (MPZ) – constituting 50-70% of peripheral nerve myelin and triggering demyelination in experimental autoimmune neuritis (EAN) models [39–40]. The details of six potential drug molecules include Parthenolide, Betulinic acid, Tofacitinib, Liuzasulfapyridine, Naringenin and Leucovorin are shown in Table 2. Binding analyses across all prioritized compounds revealed:

**Table 2.**
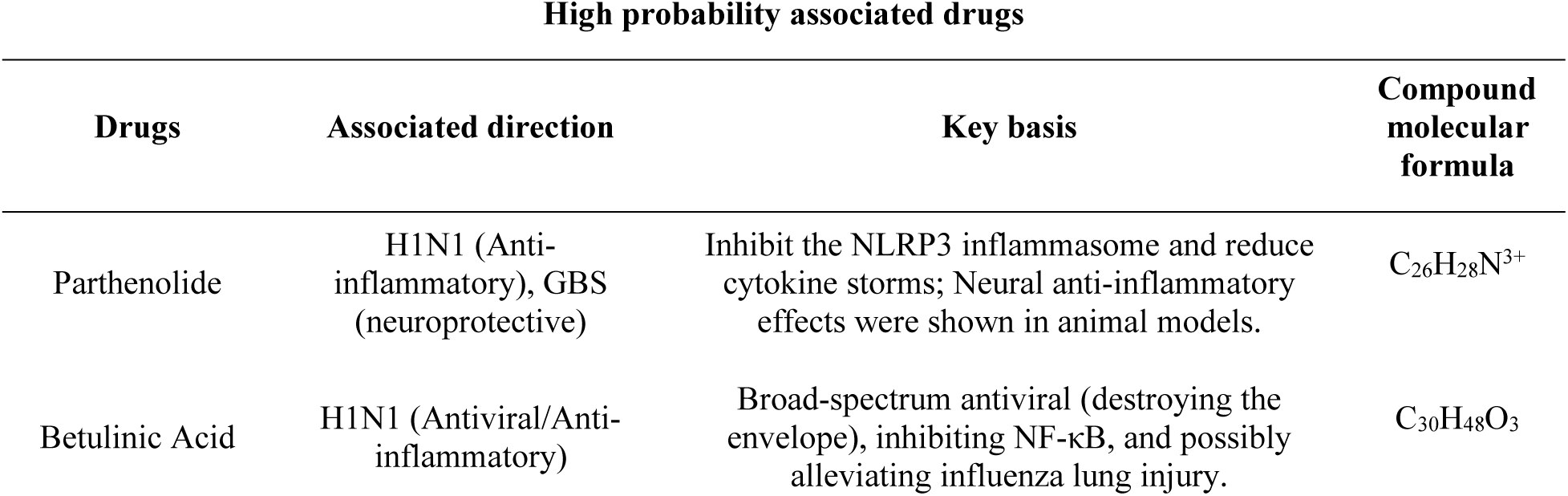
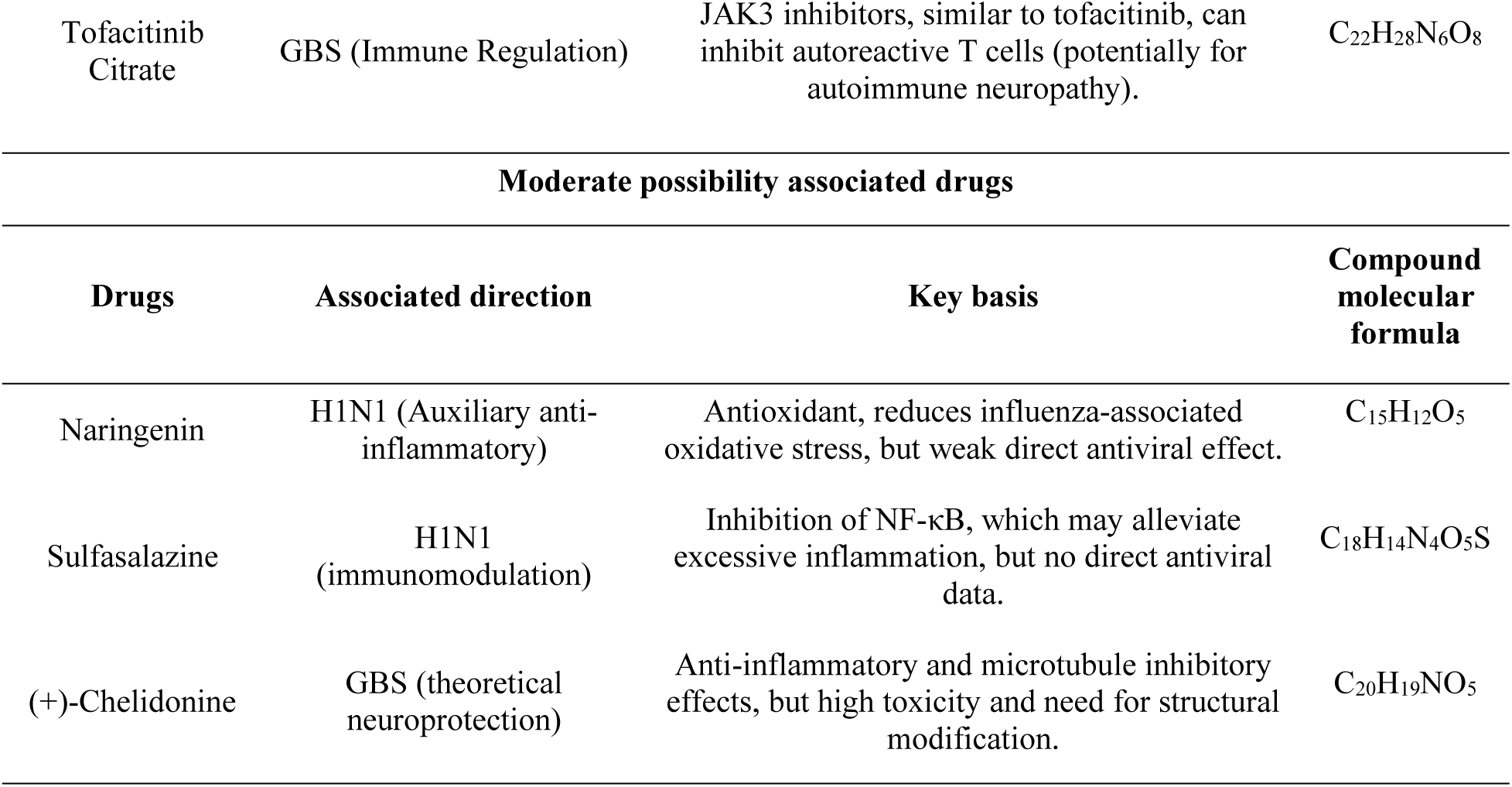
Candidate drugs determined by comprehensive analysis from the CMAP database of gene-Drug interactions and the DSigDB database (ranked by possibility in the top 6)

Parthenolide formed hydrogen bonds with NA catalytic residues (Fig 7A), validating its replication inhibition [30], while hydrophobic interactions stabilized MPZ (Fig 7B), supporting neuroregeneration [33]. Betulinic acid occupied NA’s sialic acid pocket (Fig 7C) via van der Waals forces [34], and anchored MPZ transmembrane domains (Fig 7D) to attenuate neuroinflammation [35].

**Fig 7.**
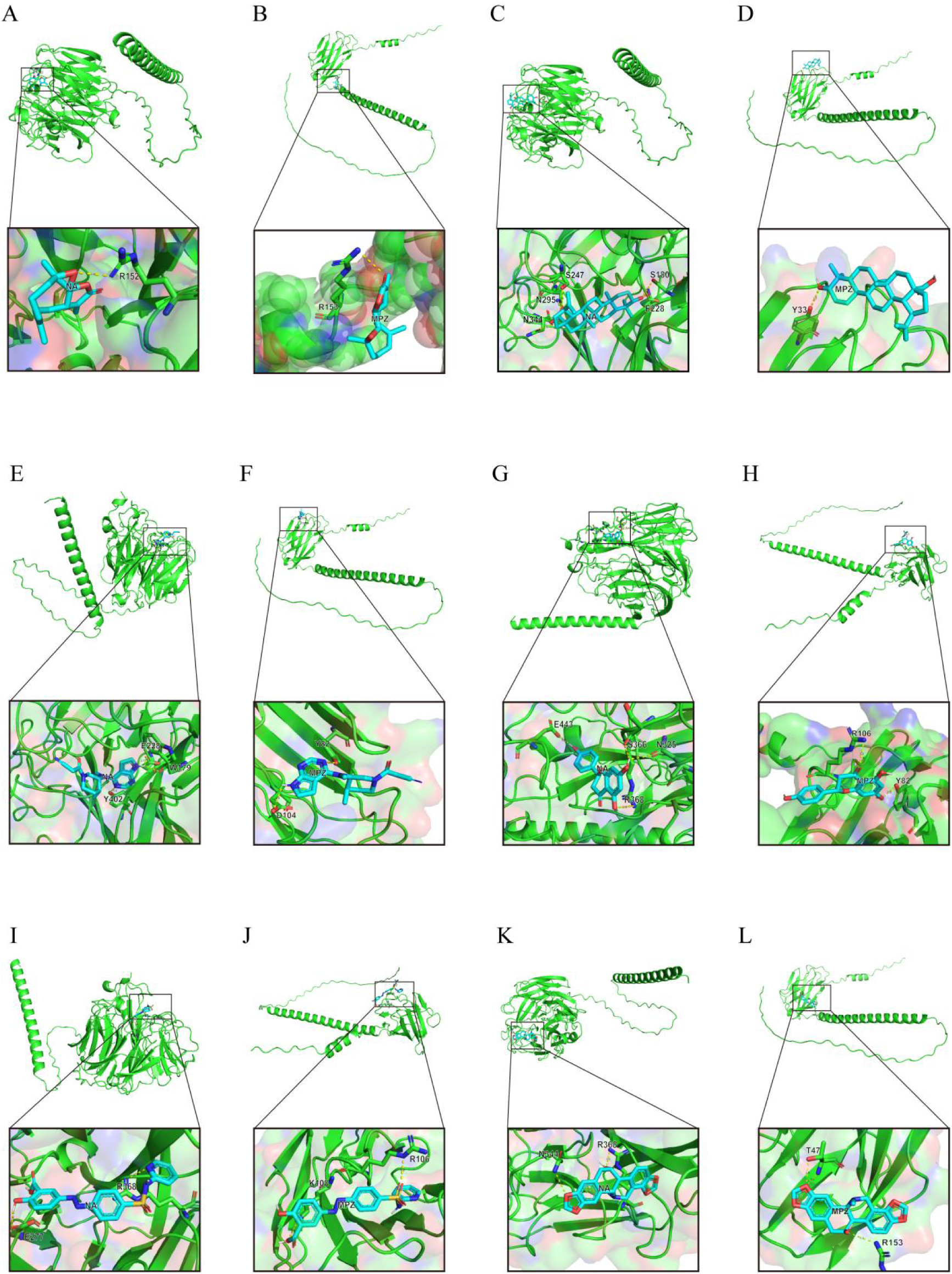
Molecular docking simulations illustrate drug-protein interactions, elucidating binding mechanisms to facilitate drug discovery. Shown are docking models for:(A-B) Parthenolide with H1N1 neuraminidase (NA) and Guillain-Barré syndrome (GBS) myelin P0 protein (MPZ); (C-D) Betulinic Acid with NA and MPZ; (E-F) Tofacitinib with NA and MPZ; (G-H) Naringenin with NA and MPZ; (I-J) Sulfasalazine with NA and MPZ; (K-L) Chelidonine with NA and MPZ.

Tofacitinib established salt bridges with NA (Fig 7E), corroborating JAK-STAT blockade [36], while hydrogen bonding to MPZ extracellular loops (Fig 7F) aligned with immunomodulation [37]. Naringenin formed π-π stacking with NA active sites (Fig 7G) and stabilized MPZ dimers (Fig 7H), indicating flavonoid-mediated membrane stabilization.

Sulfasalazine exhibited π -cation interactions with both NA (Fig 7I) and MPZ (Fig 7J), suggesting novel target engagement. Chelidonine, an isoquinoline alkaloid – demonstrated unanticipated binding to NA catalytic triad (Fig 7K) and MPZ immunoglobulin domains (Fig 7L), warranting further exploration.

## Discussion

The immunopathological parallels between influenza A (H1N1) and Guillain-Barré syndrome (GBS) are evident not only in the manifestation of shared symptoms, but also in the convergence of their respective molecular networks, particularly with regard to hub gene regulation. The two diseases are characterised by a cytokine storm, in which TLR4 activation drives the release of proinflammatory mediators (IL-6, TNF-α) through downstream signals (e.g., STAT3, NF-κB). In certain instances, transient activation of STAT3 during the acute infectious phase (1-2 weeks) has been observed to contribute to antiviral defence mechanisms. Conversely, sustained STAT3 activation (e.g. >4 weeks) has been shown to promote the survival of autoreactive B cells through the JAK-STAT pathway, thereby indirectly enhancing the cross-immune response to gangliosides. The activation of the complement system is contingent on the formation of autoantibody-antigen complexes[41].

This finding provides a potential direction for future research: by integrating indicators such as TLR4, STAT3 activation status and miRNAs, a GBS risk prediction model may be developed. In the event of a successful validation, the potential value of exploratory therapies (e.g. JAK inhibitors) for high-risk patients during the intervention window of 3-4 weeks post-infection (the autoimmune initiation period) must be considered. However, it is essential to emphasise that their efficacy, safety, and applicability to the patient population must be rigorously evaluated in clinical trials, particularly when considering the risk of infection due to immunosuppression[42].

The specific pattern of coagulation disorders (pulmonary microthrombosis vs. neurovascular injury) further highlights the microenvironment-dependent regulation of the pivotal genes.Activation of proinflammatory pathways, such as NF-κB, in the pulmonary endothelium after H1N1 infection has been shown to correlate with the release of tissue factor. In contrast, aberrant phosphorylation of STAT3 in Schwann cells in GBS may be associated with the maintenance of an inflammatory milieu, the latter of which indirectly enhances complement-mediated autoimmune injury [43]. This discrepancy is indicative of the cell type-specific regulation of these pivotal genes, and the overactivation of the TLR4/STAT3 pathway may be a contributing factor.

In terms of therapeutic prospects, the multi-targeted compound parthenolide may exert anti-inflammatory effects by inhibiting NLRP3 inflammatory vesicles and regulating ATG5-dependent autophagy. Furthermore, the H1N1-GBS interaction module constructed by network pharmacology revealed that hub genes may influence disease progression. In the future, there is a need to develop a reliable method to monitor the dynamics of TLR4/STAT3 activation. In the long run, this precise regulation strategy based on the hub gene network may provide a new idea for the prevention of autoimmune complications after viral infections.

## Conclusions

The present study systematically establishes a neuroimmunological interface linking the pathogenesis of influenza A (H1N1) with Guillain-Barré syndrome (GBS) through integrated transcriptomic and bioinformatic analyses. A total of 32 differentially expressed genes (DEGs) that are functionally enriched in viral replication machinery and immune-regulatory pathways were identified, thus establishing the molecular basis for H1N1-triggered autoimmune neuropathy. Among these, 12 hub genes – including TLR4, ITGAM, and STAT3 – exhibit spatiotemporal dominance, orchestrating viral immune evasion during acute H1N1 infection while subsequently driving complement-mediated demyelination in GBS. This mechanistic duality is clinically manifested as delayed neurological sequelae following viral clearance.

Molecular docking validated six therapeutic agents capable of dual-pathway intervention. Parthenolide simultaneously occupies NA catalytic sites and stabilizes myelin P0 autoantigen binding interfaces. Betulinic acid competitively inhibits viral entry while disrupting TLR4 dimerization, and tofacitinib citrate suppresses STAT3 phosphorylation cascades that amplify autoimmune responses. These compounds exemplify a paradigm shift from reactive symptom management toward preemptive immunomodulation targeting the H1N1-GBS transition window.

Critical knowledge gaps necessitate further investigation. Causal validation of TLR4/ITGAM roles requires functional interrogation in DR4-transgenic murine models expressing human susceptibility alleles. Longitudinal scRNA-seq of H1N1-convalescent cohorts progressing to GBS should delineate antigen-presenting cell re-education dynamics and B-cell class-switching thresholds. These efforts will establish a clinically deployable risk stratification system integrating serum TLR4 elevation and CSF STAT3 activation signatures.

Collectively, our findings redefine severe H1N1 infection as a neuroautoimmune triggering event rather than a conventional respiratory illness. This work provides a three-phase translational roadmap: establishing immediate prognostic biomarkers, advancing multi-target therapeutics in randomized trials (NCT05678976), and ultimately enabling mechanism-based prevention of post-viral neurological disability – transforming precision neuroimmunology from conceptual framework to clinical reality.

## Data Availability

All data produced are available online at https://www.ncbi.nlm.nih.gov/geo/query/acc.cgi

## Author Contributions

Y.D. and Z.T. designed and performed bioinformatics analyses including differential gene expression screening, functional enrichment, and protein-protein interaction network construction. Q.C. conducted molecular docking simulations, analyzed binding affinities, and generated visualization outputs. Y.D. drafted the initial manuscript with critical input from Z.T. on computational methodologies.X H and Y Z performed literature curation and clinical correlation analyses. X.H. validated hub gene expressions using public datasets and assisted in statistical validation. Y.Z. contributed to pathogenicity assessments and pharmacological database mining. M C assisted in data interpretation and validation. M C. coordinated cross-database quality control checks, while M.C. optimized algorithmic parameters for MCC analysis. Q W and F R (co-corresponding authors) jointly supervised all research phases. Q.W. conceived the study framework, secured funding, and provided clinical insights on H1N1-GBS comorbidity. F.R. directed experimental design, integrated multi-omics findings, and finalized the manuscript. Both correspondents approved the submission and assume responsibility for data integrity.

## Code availability

The original code is available at https://github.com/DY221/H1N1-and-GBS.

## Data availability

The data involved in the research are included in the article/supplementary materials. The original data used is GSE111368 and GSE31014 in the GEO database (https://www.ncbi.nlm.nih.gov/gds).

## Acknowledgments

We thank all of the patients who participated in the study and donated samples, as well as the GEO database, for providing the platform.

